# Immunity status of Health Care Workers post recovery from COVID-19: An online longitudinal panel survey

**DOI:** 10.1101/2020.11.27.20239426

**Authors:** S B Shah, R Chawla, A Pahade, N Bansal, A Mehta, A.K. Dewan, A Prakash, M Bhatia

## Abstract

**Background:** Corona virus has literally travelled “around the world in 80 days” akin to Fogg and Passepartoute of Jules Verne fame. Manning of corona virus disease 2019 (COVID-19) wards and ICUs, also surgery on COVID-positive patients is increasingly being relegated to that subset of health care workers (HCW) who themselves have resumed duties after surviving COVID-19 infection. Convalescent plasma therapy has been widely endorsed. Several vaccines are in the pipeline as potential preventive measures against the virus keeping HCW on the priority-list of recipients. Immunity passports are being validated for foreign travel. These events share a common presumption that exposure to COVID-19 virus (natural infection/inoculation) produces protective adaptive immunity. It is unknown whether all (COVID-19) infected patients mount a protective immune response and for how long any protective effect will last.

**Methods:** This single institutional prospective longitudinal panel survey questions were deployed to the respondents online via email/WhatsApp groups to ascertain the symptomology and immunity status of HCW in the months following COVID-19 infection. The survey was administered to the same set/cohort of health care workers over 6 months.

**Results:** 165 responses from 151 respondents (70 at 1-2months; 95 at 3-4 months including 14 at both time points) were analysed. 7.14% of infected HCW failed to develop IgG antibodies at 4-6 weeks. 91.7% HCW with IgG titres in the highest bracket had experienced anosmia. Mean antibody titres were 12.08 ± 9.56 and 9.72 ± 9.34 at 1-2 months and 3-4 months post-development of first symptom, respectively.

**Conclusion:** Understanding of COVID-19 patterns of variation in HCW may guide their deployment in the COVID ward and COVID-OTs. Revelation of this enigma (by quantification of serial IgG antibody levels) is critical for predicting response to vaccines under trial, fostering effective stratagems and tactics for pandemic control, ascertaining validity of immunity passports and understanding longevity/durability of protection by forecasting immunological memory against SARS-CoV-2.

## Introduction

Corona virus disease 2019 (COVID-19) caused by SARS-CoV-2 was declared a ‘Pandemic’ by the World Health Organization on 11 March 2020.

Manning of COVID wards and surgery on suspected/known COVID-positive patients is increasingly being relegated to health care workers (HCW) who are COVID-survivors owing to a general notion of their acquiring immunity. Convalescent plasma therapy (CPT) has been widely endorsed.^[1,2,3,4]^ However, the multicentric PLACID-trial findings report no difference in 28-day mortality between the intervention group patients (two 100ml doses of convalescent plasma) and the control group.^[2]^ It is important to gauge the existence, extent and duration of this immunity in COVID-survivors and this is the research question we seek to answer. Contrary to the assumption of acquiring immunity, patients getting re-infected within a short span of recovery have also been reported.^[5,6].^ Thus, the immune response following COVID-19 infection is unclear. This survey aims to determine whether the severity, duration or type of symptoms affect the development of immunity as measured by the antibody titre. It also seeks to know if the antibody titres remain constant or decline over six months and by how much.

As per quantitative antibody tests, positive results for IgG, IgM, and IgA are generally described as > 1.0 arbitrary unit (AU)/mL. Values for IgG antibodies > 6.5 AU/mL correspond to an antibody titer of approximately > 1:320 and IgG values > 20 AU/mL correspond to titers > 1:1000. An IgG antibody titer > 1:160 has been recommended by the Food and Drug Administration (FDA) as a threshold for suitability of convalescent plasma for donation as CPT.^[7]^

To analyze long-term immunity, it’s imperative that serological tests focus on long-lived, extremely-specific IgG isotype and not merely the shorter-acting, less specific and pro-inflammatory IgM antibody.^[8]^ Other defense mechanisms like formation of B memory cells, and T-cell immunity cannot be assessed by serological tests.^[4,8]^ Besides, the mere presence of IgG antibodies does not guarantee protection from future infection^.[9]^ No re-infection of rhesus macaques re-exposed to SARS-CoV-2 4weeks after initial infection was reported.^[10]^ A non-peer-reviewed study reports robust, year-long immunity in COVID-19 survivors raising optimism that production of IgG maybe protective against SARS-CoV-2. Despite paucity of data and lack of definitive evidence, vaccine trials are underway. Nobody knows how long immunity will persist if it actually exists.

Existing body of evidence maintains that IgG-antibodies against coronavirus family (cCoVs) peak around 2 weeks post-infection and revert to baseline values after one year. Three out of four cCoVs have been implicated in reinfections whose basis is poorly understood.^[11]^. Regression of protective immunity and re-exposure to mutant strains of the virus may explain this phenomenon. SARS-CoV antibodies peak approximately 3-4 months post infection and progressively become undetectable by 6 years post-exposure.^[12]^ Neutralizing antibodies may persist even 3 years post-exposure to MERS-CoV virus.^[13]^

**Our primary objective was to** find out if a correlation exists between the symptomology (type, severity and duration of symptoms) and antibody (IgG) titre post recovery from COVID-19.

**Our secondary objectives** are to find out if the IgG titre falls with time (1-2months, 3-4months and six months) and by how much.

## Methodology

This prospective longitudinal panel survey was carried out after written informed consent from all patients, and approval from the Scientific Committee and Institutional Review Board. The survey is being administered to the same set/cohort of HCW spanning six months. The survey questionnaire comprising multiple-choice, dichotomous, matrix and Likert-scale questions was deployed to the respondents online via email/WhatsApp.

An online survey software (Google forms), was utilized to deploy the survey questions and get analyzed data on a dashboard which keeps updating real-time as respondents partake the online survey. Data presentation on this dashboard comprises charts and graphs for the ease of statistical analysis (figure-1).

**Figure-1:**
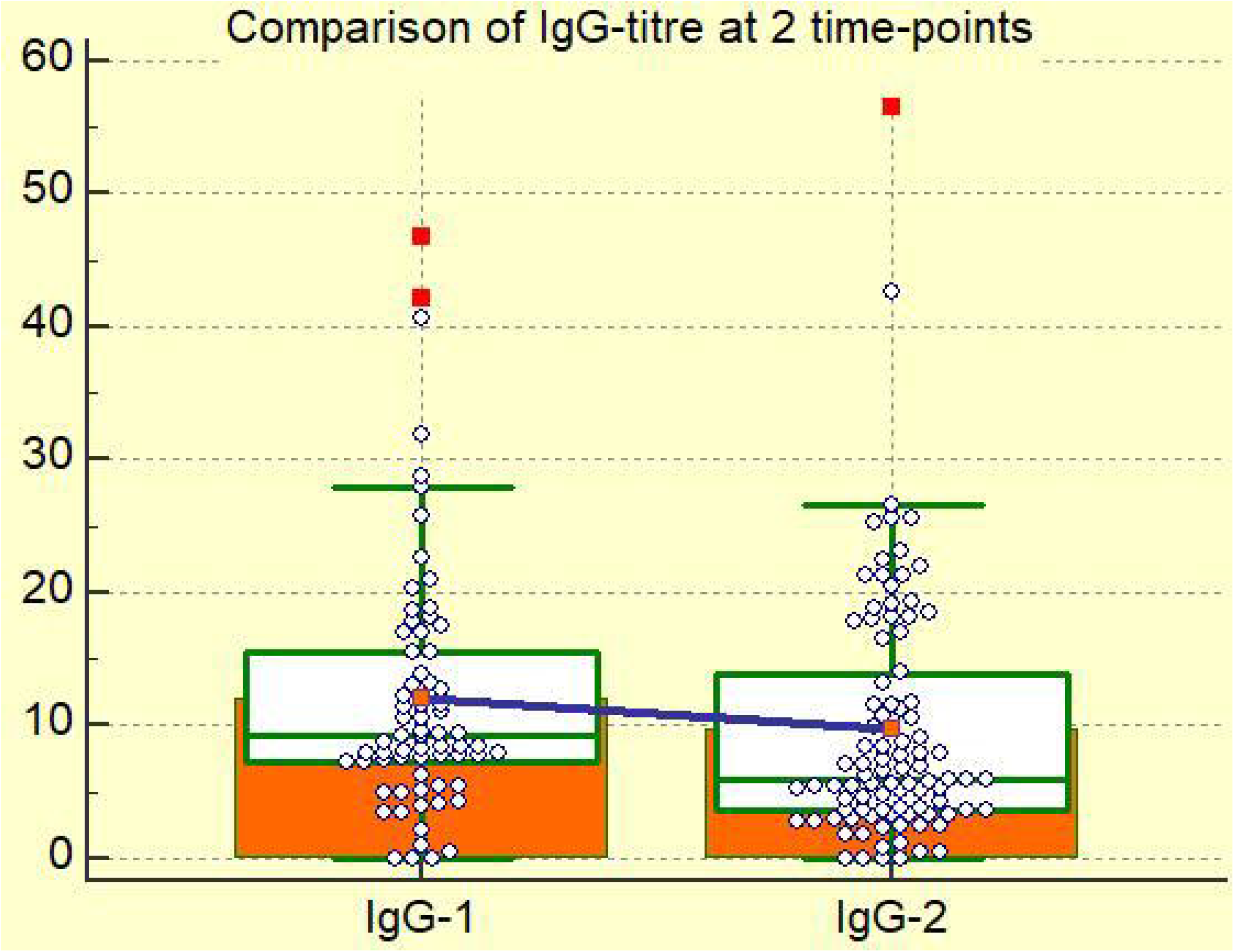
Symptomology of respondents: type and duration of symptoms

Initial three steps of the survey (defining the population and sample, deciding the type of survey, designing the survey-questionnaire) were completed before and the remaining three steps (distribution of survey and response-collection, survey-result analysis, penning the survey results) were conducted after ethics committee approval.

All HCW, employed at Rajiv Gandhi Cancer Institute and Research Centre (RGCIRC) with a history of being a laboratory confirmed COVID-positive patient and with antibody tests conducted on them were included in the study. Non-HCW and HCW without Reverse transcription polymerase chain reaction (RT-PCR)/Gene Xpert reports were excluded from the survey.

Antibodies binding to the receptor binding domain of the surface glycoprotein/spike (S) protein of SARS-CoV-2 can neutralize the virus.^[6,14,15]^ The antibody test kit utilized at RGCIRC (VITROS Immunodiagnostic Products Anti-SARS-CoV-2 IgG) is based on the high throughput automated chemiluminescence immunoassay (CLIA) technology and the antibodies tested are those produced against the S-protein of SARS-CoV-2. It is an immunometric test utilizing ECi/ECiQ, 3600, 5600/XT 7600 system with incubation time 37mins, time to first result 48mins and an intravenous serum sample of 20 μL tested at 37 °C. Positive Percent Agreement to PCR of 90.0% and 100% clinical specificity (95% CI: 99.1–100.0%) are additional features.^[16]^

It describes values<1 as non-reactive, those between 1-1.46 as providing low level of immunity, those between 1.46-18.45 as medium levels of protection and values above 18.45 as providing high levels of protection.

Data collection is being done at three time points. T1, T2 and T3 at 1-2 months, 3-4 months and 6 months post-development of first COVID-19 symptom/positive RTPCR test whichever is earlier respectively, from the same cohort of HCW.

Data analysis shall be dual phased. Phase I comprises analysis of first responses from first 220 HCW who respond after getting their IgG antibody tests done. Phase II comprises analysis of 2^nd^ and 3^rd^ responses from the same cohort of HCW and corelating them with the 1st response.

This manuscript comprises an interim analysis (phase I) of early results from 1^st^ 151 HCW. To adequately power the survey a sample-size of 200 was arrived at. The target number was increased to 220 allowing for 10% dropouts. First responses of first 151 HCW fitting the inclusion criteria have been analyzed in this interim analysis since important trends have appeared.

### Privacy

To ensure confidentiality, identification numbers were assigned to each participant. The list of numbers assigned to names was kept separate from all other surveyed information, and only the identification numbers kept with the collected data, so that the list of names and identification numbers can subsequently be used to enable linking of survey data with archival records.

### Ethical Implications

The survey was conducted entirely on a voluntary basis without any binding on the respondents to attempt all questions

### Statistical Analysis

Categorical data has been expressed as absolute numbers and percentages. Continuous data is presented as mean with 95% confidence intervals, standard deviation, and box-whisker plots. Kolmogorov-Smirnov test was used to ascertain normal distribution of data. A non-parametric test (Wilcoxon-Mann-Whitney test) was used to determine whether survey responses to two different questions are statistically related. A p-value <0.05 was considered statistically significant. MedCalc statistical software (version 15; MedCalc Software Ltd; Ostend, Belgium) was utilized for statistical analysis.

## Results

The first 151 responses meeting inclusion criteria were analyzed. 120/151 (79.47%) belonged to 20-40y age group, 25/151(16.56%) in the 40-60y age bracket, 4/151 (2.65%) in more than 60y bracket and 2/151 (1.32%) in 20y bracket.

95/151 (62.91%) of the COVID-19 infected HCW are females and 56 (37.09%) are male.

81/151 (53.64%) respondents are nurses, 26/151(17.21%) are doctors, 20/151 (13.25%) from hospital administration, 10/151 (6.62%) are OT/radiology/lab technicians, 9/151 (5.96%) are information-technology staff and 5/151 (3.31%) are pharmacists.

145 respondents communicated in English (96.02%) and six in Hindi (3.97%).

32 and 3 respondents respectively, have a blood group of A-positive (21.2%) and A-negative (1.99%) respectively. 51 respondents are B-positive (33.77%) while none are B-negative (0). 15 and 2 respondents have an AB positive (9.93%) and AB negative (1.32%) blood group respectively. 46 and 2 respondents respectively have an O-positive (30.46%) and O-negative (1.32%) blood group respectively.

75/151 (49.67%) of HCW do not wear spectacles, 54/151 (35.76%) wear spectacles, 21/151 (13.90%) wear them occasionally.

114/151 (75.49%) respondents had no co-morbidity whatsoever, 37 had one or more co-morbidity (hypertension (11(7.28%)), obesity (12(7.94%)), diabetes (4(2.64%)), asthma (6(3.97%)), hypothyroidism (53.31%)), hyperthyroidism (1(0.66%)) PCOD (1(0.66%)) GERD (1(0.66%)), elevated cholesterol (1(0.66%)))

The symptomology (fever, cough, sore-throat, dyspnoea, post-nasal drip, retro-orbital pain/headache, calf-pain, myalgia/bodyache, malaise/weakness, diarrhoea, anosmia, loss of taste, COVID-toes) with duration is depicted by the bar chart (Figure-1). Maculopapular rash (4), hair loss (3), vertigo, severe backache and deranged lipid profile in (2-each), chest pain, joint pain, severe neck pain radiating to left arm, tingling-numbness of toes, pricking pain in throat, itching, constipation, severe shivering followed by profuse sweating, lack of concentration and somnolence (1-each) were additional symptoms observed. One individual (normotensive and euthyroid earlier) developed hypertension and extremely elevated TSH-levels (156 units). Re-appearance of symptoms after 5 weeks was observed in another respondent.

Based on their symptom-complex 95 (62.91%) respondents were categorized as mild, 45 (29.80%) moderate and 11 (7.28%) as severe infections. There was one death that occurred before the antibody titre could be done (hence excluded from the analysis).This 48y old male HCW was a well-controlled diabetic who underwent dialysis at RGCI for COVID-induced deranged KFT and septic shock as a last resort (Cycle threshold value 12.86 by RT-PCR; D-dimer level 9634ng/ml; CRP 4.8mg%;IL-6 48.6pg/ml; procalcitonin 4.59ng/ml; hemoglobin 7.9gm; TLC 35460/ml; platelet count 60000/ml)

### Fever

89/151(58.94%) HCW developed fever lasting 1-2 days, 31/151(20.53%) for 3-7 days, 9/151(5.96%) for 1-2w and 2/151(1.32%) for >2 weeks. 20/151(13.25%) HCW remained afebrile. 22/151(14.57%) respondents developed no fever/<99°F, whereas 36/151(23.84%) developed mild fever (98.6-100°F), 35/151(23.18%) had 100-101F fever, 21/151(13.91%) had 101-102F, 14/151(9.27%) developed 102-103F, 17/151(13.36%) had 103-104 F and 6/151(3.97%) had >104 F body temperature, respectively.

### Breathing difficulty

26/151(17.22%) HCW developed dyspnea/breathing difficulty lasting 1-2 days, 19/151(12.58%) for 3-7d, 6/151 (3.97%) for 1-2weeks and 5/151(3.31%) for >2weeks while 95/151(62.91%) HCW experienced no dyspnea/breathing difficulty.

### Anosmia

35/151(23.18%) HCW developed anosmia lasting 1-2 days, 26/151(17.22%) HCW for 3-7 days, 16/151(10.6%) for 1-2weeks, 9/151(5.96%) for >2weeks while in 65/151 (42.2%) HCW, the sense of smell was unaffected.

### Quarantine Management

87/151 (57.62%) of respondents were home-quarantined, two of whom were shifted to RGCI COVID-ward when their condition deteriorated at home. 59/148 (39.07%) were quarantined at RGCI and remaining 5/148(3.38%) were quarantined at a government-designated isolation centre/corporate hospital.

**Treatment** included CPT 3/151(1.99%), remdesivir 6/151 (3.97%), hydroxychloroquine 43/151(28.48%), alternate medicine 60/151(39.74%), clexane 9/151(5.96%), steroids (13/151(8.61%)), and azithromycin 62/151(41.06%) in varying combinations besides observation, monitoring and paracetamol.

**Long-COVID symptoms** included chronic fatigue 50/151(33.11%), breathlessness 20/151(13.25%), bodyache7/151(4.64%), headache5/151(3.31%), myalgia, joint pain, lower limb pain and gastritis (3-each), deranged lipid profile and itching (2-each) and weakening of eyesight, hypertension, hypothyroidism, deranged liver function tests, chest tightening, loss of concentrating power, persisting anosmia, diarrhoea (1-each). 73/151 (48.34%) HCW did not suffer from long COVID symptoms

### Immunoglobin assay

70/151(46.35%) RT-PCR COVID-19 positive HCW were tested for IgG at 1-2m post-development of first symptom while 95/151(62.91%) HCW were tested at 3-4months (figure-2). 12/151(7.94%) respondents had an antibody titre <1(5/151(3.31%) tested at 4-6weeks and 7(4.63%) tested at 3-4 months) The mean IgG-antibody titre at 1-2 months post onset of symptoms (POS) was 12.08 (95% CI for the mean being 9.80 to 14.36 and standard deviation (SD) being 9.59), the median value was 9.24, while the lowest and highest values were 0.00 and 46.8 respectively. Similarly, at 3-4 months, mean ± SD was 9.72 ± 9.34 (Table-1).

**Table-1:** Summary statistics

| Parameter | Ab titre at 1-2m | Ab titre at 3-4m |
| --- | --- | --- |
| Sample size | 70 | 95 |
| Lowest value | 0.00 | 0.00 |
| Highest value | <u>46.80</u> | <u>56.50</u> |
| Arithmetic mean | 12.08 | 9.72 |
| 95% CI for the mean | 9.80 to 14.36 | 7.81 to 11.62 |
| Median | 9.24 | 6.02 |
| 95% CI for the median | 8.00 to 11.47 | 5.32 to 7.89 |
| Variance | 91.37 | 87.25 |
| Standard deviation | 9.59 | 9.34 |
| Relative standard deviation | 0.7911 (79.11%) | 0.9612 (96.12%) |
| Standard error of the mean | 1.14 | 0.96 |
| Coefficient of Skewness | 1.71 (P<0.0001) | 2.11 (P<0.0001) |
| Coefficient of Kurtosis | 3.39 (P=0.0014) | 6.6739 (P<0.0001) |
| Kolmogorov-Smirnov test<sup>a</sup><br>for Normal distribution | D=0.1677<br>reject Normality (P<0.0001) | D=0.1877<br>reject Normality (P<0.0001) |

**Figure-2:**
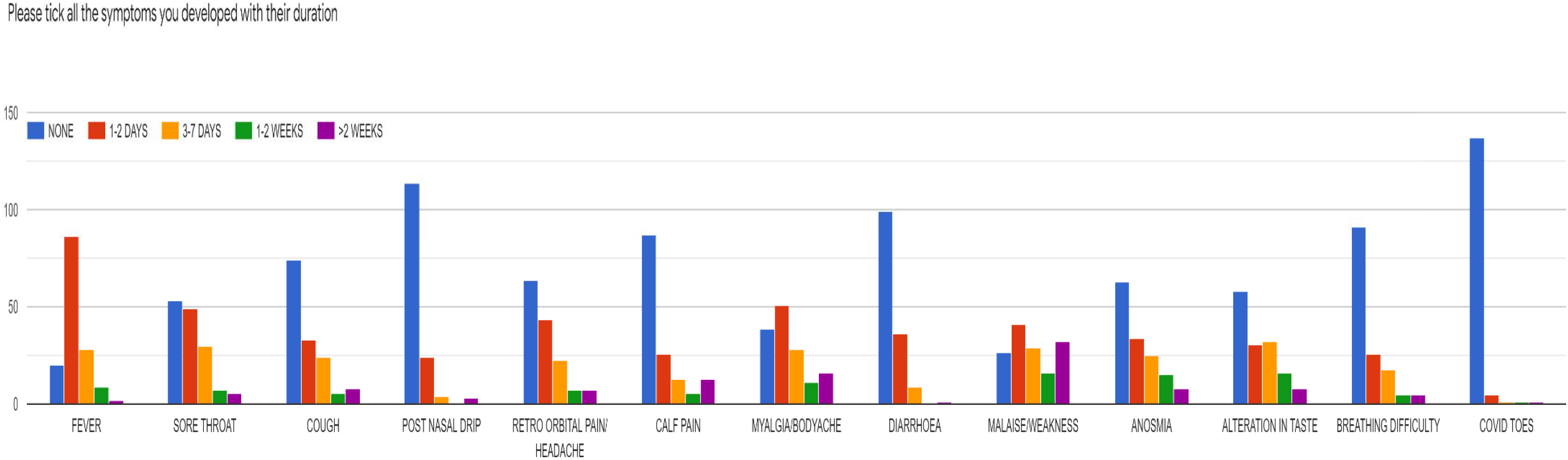
Comparison of IgG titres at two different time points

5/70 (7.14%) HCW had an antibody titre <1 at 4-6w POS. Clinical features of these HCW with no antibodies detectable at 1-2months have been tabulated (Table-2).

**Table-2:** Clinical features of HCW with no antibodies detectable at 1-2months post first symptom

| Clinical Features | HCW-1 | HCW-2 | HCW-3 | HCW-4 | HCW-5 |
| --- | --- | --- | --- | --- | --- |
| IgG Ab titre | 0.00 | 0.01 | 0.01 | 0.94 | 0.57 |
| Age | 40-60 | 20-40 | 40-60 | 20-40 | 40-60 |
| Sex | F | M | F | M | F |
| Blood Group | O+ | O+ | O+ | O+ | B+ |
| Co-Morbidity | HTN | Nil | Nil | Nil | Nil |
| Fever | 1-2w<br>101-102°F | 3-7d<br>>104°F | Nil | Nil | 3-7d<br>101-102°F |
| Sore throat | 3-7d | Nil | Nil | 1-2d | 3-7d |
| Post nasal drip | Nil | Nil | 1-2d | Nil | Nil |
| Headache | >2w | Nil | Nil | 1-2d | 3-7d |
| Calf pain | >2w | 3-7d | Nil | Nil | 1-2w |
| Myalgia/Bodyache | >2w | 3-7d | 3-7d | 1-2d | 1-2w |
| Malaise/weakness | >2w | 3-7d | 1-2d | 3-7d | >2w |
| Diarrhoea | 3-7d | Nil | Nil | Nil | 3-7d |
| Anosmia | Nil | Nil | Nil | 1-2w | 1-2d |
| Loss of taste | Nil | Nil | 1-2w | 1-2w | 1-2d |
| Dyspnoea | Nil | Nil | Nil | 1-2d | Nil |
| COVID-toes | Nil | Nil | Nil | 1-2d | Nil |
| Treatment | Obs/Monit<br>PCM<br>Azithromycin<br>Alt Med<br>Lomofen | Obs/Monit<br>PCM<br>HCQ<br>Alt Med<br>Vit-C | Obs/Monit<br>Clexane<br>HCQ<br>Alt Med<br>Vit-C<br>Multivit | Obs/Monit<br>PCM<br>Azithromycin<br>Alt Med<br>Vit-C Vit-B<br>Pantoprazole | Obs/Monit<br>PCM<br>Azithromycin<br>Alt Med |
| Long COVID | Bodyache<br>Rt knee pain | Nil | Chronic<br>fatigue | Nil | Chronic<br>fatigue |

6/70(8.57%) had low level of protective antibodies, 47/70(67.14%) had medium level and 12/70(17.14%) had high levels of protective antibodies. Hence majority of HCW developed medium to high levels of IgG.

12/70(17.14%) HCW developed IgG titre > 18.45 at 1-2m POS, synonymous with high degree of protection. Their symptomology is tabulated below (Table-3)

**Table-3:**
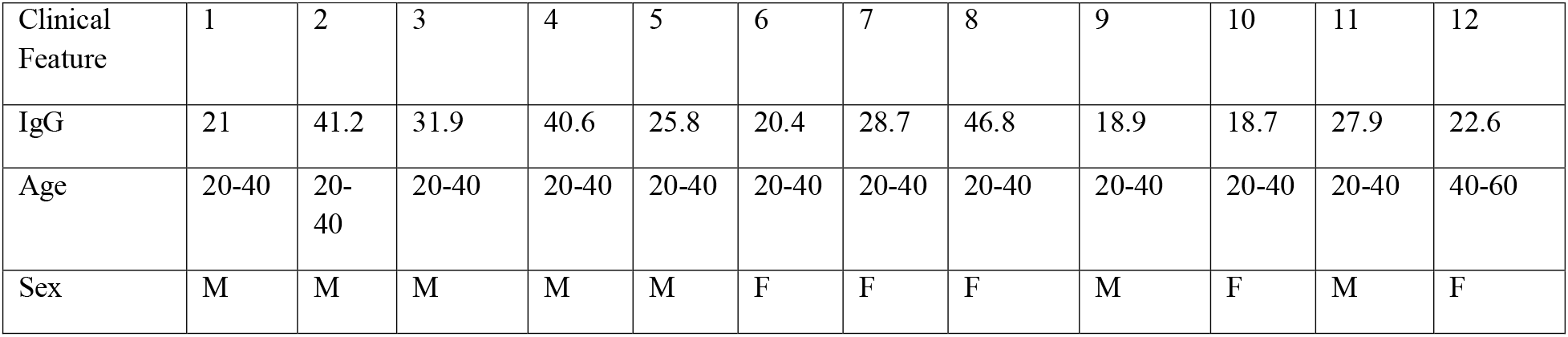
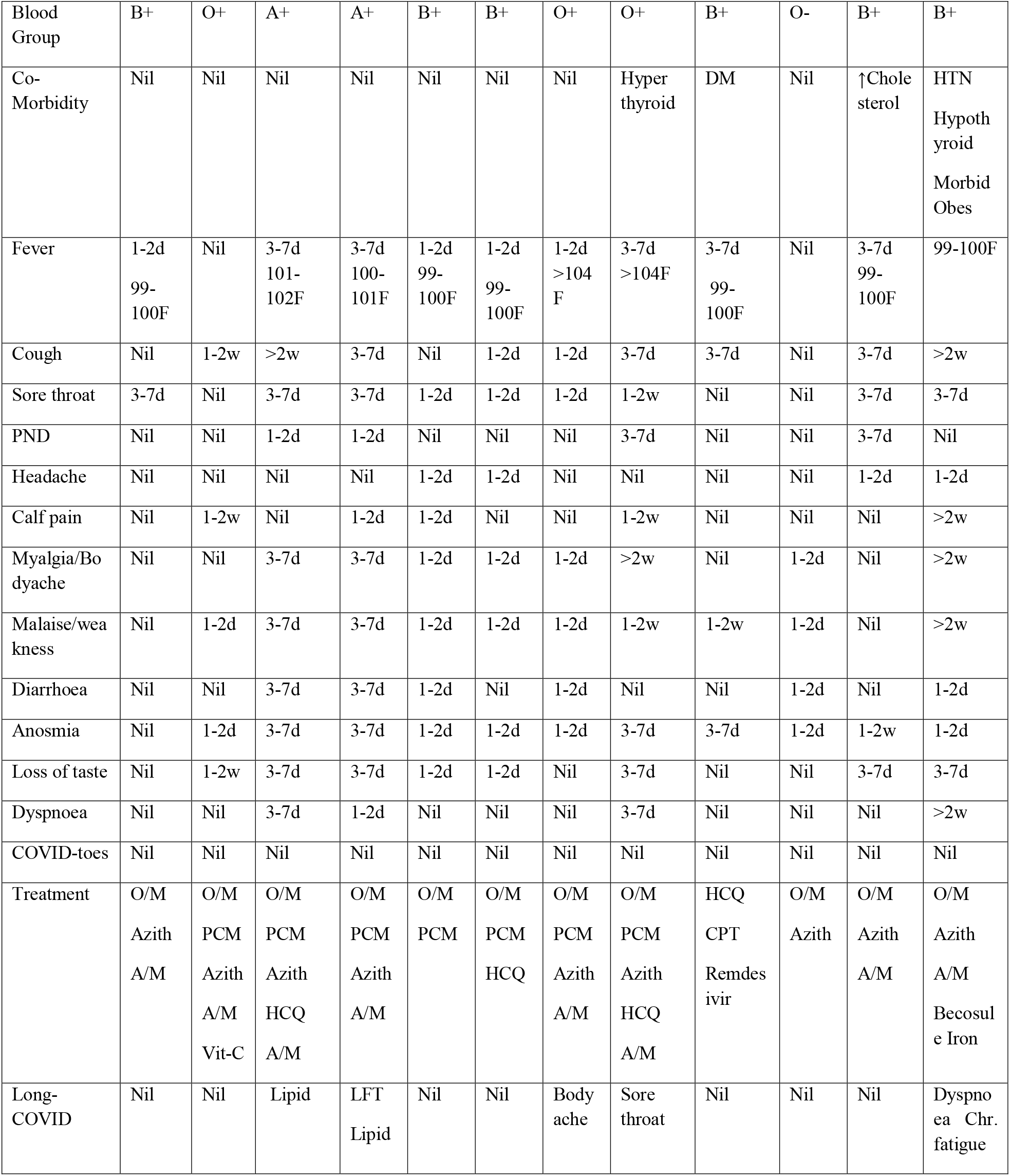
Clinical features of HCW with maximum antibodies detected at 1-2months post first symptom

**Table-4:**
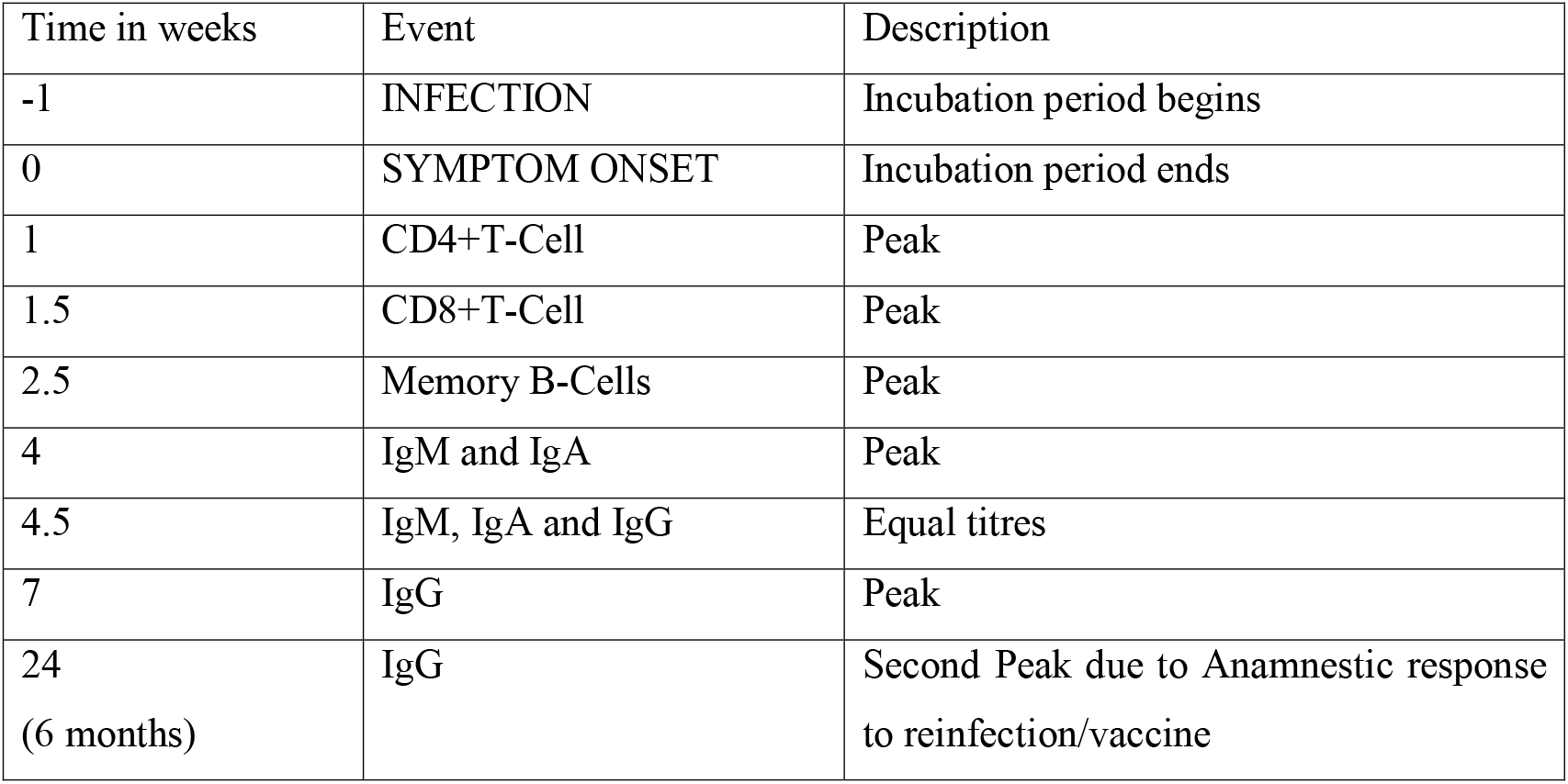
Adaptive immune response to COVID-19 infection: How valid is the immunity passport?

| Time in weeks | Event | Description |
| --- | --- | --- |
| -1 | INFECTION | Incubation period begins |
| 0 | SYMPTOM ONSET | Incubation period ends |
| 1 | CD4+T-Cell | Peak |
| 1.5 | CD8+T-Cell | Peak |
| 2.5 | Memory B-Cells | Peak |
| 4 | IgM and IgA | Peak |
| 4.5 | IgM, IgA and IgG | Equal titres |
| 7 | IgG | Peak |
| 24<br>(6 months) | IgG | Second Peak due to Anamnestic response to reinfection/vaccine |

95/151(62.91%) HCW were tested at 3-4m. The mean serum IgG-titres at 3-4m POS was 9.7 (ranged from 0 to 56.5). Out of these, 7/95(7.36%) had a titre<1, 28/95(29.47%) had low levels of protective IgG, 44/95(46.31%) had medium levels of protective IgG and 16/95(16.84%) HCW had an antibody titre in the highest bracket at 3-4m. Excluding outliers (far-out for 1-2m: 42.2, 46.8; far out for 3-4m: 56.5) the mean values were 11.13±7.85 and 9.22± 8.03 for IgG-titre at 1-2m and 3-4m respectively.

14/151 (9.27%) HCW got their antibody titres tested at 4-6 weeks POS and again at 3-3.5 months POS. There was a fall in IgG-titre during this period in 9 (64.3%), no change in one (7.1%) and a rise in IgG-titre in four (28.6%) HCW. All were asymptomatic after initial recovery.

## Discussion

The difference in denominators between our hospital HCW and the general population is largely responsible for the discrepancy in the demographic parameters of the COVID-affected HCW and the general trend. Our hospital record shows total 1482 HCW out of which 294 have been infected by COVID-19 from May to October 2020 (prevalence 21.1%). 16 HCW have left the institution post-recovery from COVID-19 in past 5 months, the attrition attributable to completion of DNB tenure (4), non-resumption of duties post-COVID (7), personal issues (marriage, retirement, lucrative job in native place) and one COVID-death.

Less than 14% of COVID-19 infections in India afflict the 20-40 years age-group.^[17]^ In contrast, 80% of our affected respondents fell in this age-bracket. Age-distribution of our HCW (<20y=2, 20-40y =1161, 40-60y = 306; >60 years =13) shows that majority (78%) belong to 20-40y age-bracket, which may account for the majority of COVID-affected respondents (80%) being in the 20-40y age-group. 0.88% of total HCW are above 60y of age but 2.65% of COVID-affected HCW fall in this bracket, also, the single mortality belonging to the 40-60y age group, emphasizes that incidence and gravity of COVID-19 is higher in the older population, which is consistent with other studies.^[16]^

Contrary to the existing data suggesting that both the incidence and severity of COVID-19 infection is higher in male gender,^[18,19]^ in our survey, 63% were females because majority of HCW (871/1482;58.8%) at RGCI are females. The nursing staff are predominantly females. So, if a smaller number of males are exposed then proportionately lesser number are likely to get infected. Incidentally, the nursing staff are hostel dwellers and constitute a social bubble of sorts. Lack of social distancing within this bubble predisposes all of them to catch the infection even if a single HCW develops it. Nearly half of those infected belonging to the nursing staff is once again attributable to the social bubble reason cited above. Despite this, the more severe cases and the solitary death were of male gender.

The maximum number of respondents belonged to B-positive blood group. All blood groups except B-negative were affected. This could either be purely coincidental or this blood group might confer some degree of immunity to the virus. Contrarily, as per a hypothesis propounded by Silva-Filho et al,^[20]^ blood group-A is most susceptible and blood Group-O is least susceptible to COVID-19 infection.

Nearly 65% of respondents not wearing spectacles, highlights the fact that spectacles might have some protective effect by acting as a barrier to transmission of the virus.^[21]^

The recovery rate was 99.7% in our institution with one mortality. This is better than the global recovery rate of 96-97% and Indian recovery rate of 98-99%.^[22,23]^ Explanations abound. Firstly, because the cohort of HCW is of a younger age group. Secondly, all of them have received BCG vaccination at birth.^[24]^ Thirdly, HCQ-prophylaxis^[24]^ was offered free of cost by the hospital to all its HCW and finally face-mask etiquettes, social distancing and repeated hand washing were adhered to as per the institutional protocol which could have reduced the viral load. Early diagnosis and easy access to medical support ensuring timely medical intervention was attributable to them being HCWs, and the hospital providing robust facilities for its COVID-infected employees.1-2 months was chosen as the timing of the 1^st^ IgG antibody test because IgG levels are known to peak at 7 weeks (Table-3).^[25]^

Not much is known as to the level at which antibody titres stabilize after recovery. Post COVID-19 acute thyroiditis is an interesting finding reported, that needs to be explored further.

Interestingly, this male HCW when testing at 3 months post-infection, developed an IgG-titre of 23.2AU at 3months. His peak IgG was presumably higher.

Home quarantine sufficed in more than half the respondents and seems to be a good option in HCW with mild symptoms.

Out of the 70 HCW tested at 1-2m, 5 HCW, failed to develop any antibodies despite developing symptoms (high fever (>101°F in two and >104°F) myalgia and malaise/weakness). Notably, all except one had an O+ blood group which may implicate this blood group in developing low antibody titre. All five had consumed immunity boosting alternative medicine which questions its efficacy in building immunity. All three females but none of the males developed long-COVID (chronic fatigue) which could be attributable to the older age bracket(40-60y) of females versus males (20-40y). Another 8.6% of HCW (with low IgG-titres) may not only have poor protection according to the multiple-hit model of neutralization but may also suffer from antibody-dependent enhancement phenomenon.^[6]^

11/12 (91.7%) HCW with antibody titre in the highest bracket (>18.5) belonged to the 20-40y age group and the same percentage experienced anosmia of varying duration, which implies that both younger age group and anosmia produce a higher protective antibody titre. The HCW with multiple co-morbidities (morbid obesity, hypertension, hypothyroidism) still has dyspnoea and chronic fatigue 3m post-infection despite a high antibody-titre.

The mean IgG-titre noted at 3-4m was 2.36AU less than that recorded at 1-2m (1.91AU after excluding far-outliers from the analysis), pointing towards a gradual decline in antibody titre with time. This decline is not clinically significant, since both 12.08 and 9.72 lie in the moderate immunity bracket. This implies that despite a decline, the average HCW still harbored protective levels of antibodies and could be deployed in frontline management of COVID patients, provided he/she was amongst those who developed antibodies and his/her individual titres were not in the low level of protection bracket (15.7% HCW at 1-2m; 38.8% HCW at 3-4m). 4/14 HCW showing a rise in IgG titre at 3-4 months as compared to 1-2months could be attributed to asymptomatic reinfection by a mutant strain. Our HCW cohort displayed greater longevity and gentler decline in antibody titres as compared to general population^[26-29]^ probably due to repeated exposure to low viral loads (despite PPE and precautions) as a professional hazard, akin to booster doses of vaccination or even an anamnestic response.

This could also be extrapolated to convalescent plasma donation and immunity passports. The PLACID-trial reports no difference in development of severe disease (PaO2/FiO2<100mmHg) or 28-day mortality between the intervention group patients (two 100ml doses of convalescent plasma) and the control group whereas Cao et al report its usefulness.^[3,1]^ Stringent criteria for plasma donation like elapsing of minimum 28days post-recovery from symptoms, minimum 14 days post negative RT-PCR report, age 18-60 years, body weight >50kg, absence of comorbidity (hepatorenal; cardiopulmonary; uncontrolled diabetes; hypertension), nulliparity (more than half the women HCW have been pregnant earlier), would leave still fewer donors ideally suitable for CPT.^[4]^ Donor-selection variations could explain the contradictory results of clinical trials and case-series on usefulness of CPT.^[1,2,3]^

But an unanswered question remains as to whether asymptomatic HCW with moderate levels of IgG, who simultaneously test positive for SARS-COV-2 by the RT-PCR test are non-infective to others. Potential vaccines may also be expected to produce an IgG titre of 12.08 at 1-2m and 9.72 at 3-4 months and are expected to provide reasonable protection for at least 4 months. The picture at 6 months is yet to unfold, but appears to be encouraging. The corollary is that 7.14% of population (corresponding to that subset of HCW who did not produce adaptive IgG antibody 1-2m post-infection) may not develop any antibodies in response to a potential vaccine. Several vaccines claim efficacy in 95% of vaccine-recipients as per ongoing trials. These figures seem slightly inflated even in context of stringent clinical trials milieu. Interruptions in cold-storage chains once the vaccines are available to the general public seem unavoidable considering that these vaccines require storage temperatures between minus 70°C and minus 20°C.^[30]^ Also, immunity develops only 4weeks after the initial vaccine shot of the dual-dose phased vaccination schedule which is consistent with our result of high IgG titre at 1-2months following exposure.

### Strengths of this survey

Firstly, uniformity provided by a single kit utilized to test IgG levels in all the HCW. Secondly, the institutional laboratory provides uniform testing conditions (right from blood sample collection to analysis of samples to generate reports) for all samples and all tests were conducted by the same team of medical/paramedical staff eliminating instrumental and observer bias. Thirdly, all blood samples have been stored in the hospital blood bank repository with a provision of re-running them in future if required. The same cohort of HCW can be followed up infinitely, till end of the pandemic/development of a vaccine.

### Limitations of the survey

Many HCW who tested COVID-positive by the RT-PCR test between May-July 2020 (prior to initiation of this survey) did not undergo antibody tests at 1-2 months post development of symptoms and did so only at 3-4 months post-infection. This is attributable to various reasons (initial non-availability of IgG antibody testing facility in Delhi as per Delhi government policy, lack of awareness about the test, lack of hospital policy leading to lack of prescription of this test, lack of subsidy on conduction of the test). Many HCW who tested positive did not get their antibody test done (were not convinced of its usefulness, did not find time due to shift duties and quarantine policies in COVID-ward and ICU, survey questionnaire in spam folder of email, attrition from institution, believed they might be obliged to donate convalescent plasma if their IgG-titre was found to be high etc)

## Conclusion

This survey has shed light on the immunity status of HCW in the months following COVID-19 infection. Findings from the present study shall contribute to understanding of COVID-19 patterns of variation in HCW and guide their deployment in the COVID-areas (wards; ICUs; OTs). Convalescent plasma IgG levels are labile and around 40% of patients may not qualify as donors at 3-4 months post development of the first COVID-symptom. Vaccines may not give protection in at least 7.14% of recipients.

## Data Availability

All data referred to in the manuscript is available

## Acknowledgements

We deeply acknowledge with gratitude the help and support provided by Dr S.K.Rawal (M.D.), Ms. Rekha (H.R. Department), Mr. Hemant (blood sample collection) and Dr Laxmi (Casualty) and all the HCW of RGCI who are COVID-survivors.

## Survey Questionnaire

Attached

